# Factors Associated with Smartphone Addiction among Students of Islamic University in Uganda: A Cross-Sectional Study

**DOI:** 10.64898/2026.05.07.26352672

**Authors:** Mayanja Akram Mukalazi, Aremu Abdulmujeeb Babatunde

## Abstract

**Background:** Smartphone addiction is an emerging public health concern among university students in sub-Saharan Africa. Limited data exist on its prevalence and associated factors in Uganda.

**Objective:** This study aimed to determine the prevalence of smartphone addiction and its associated sociodemographic and economic factors among students at Islamic University in Uganda (IUIU).

**Methods:** A cross-sectional study was conducted among 287 undergraduate students at IUIU Kampala campus. Data were collected using a structured self-administered questionnaire incorporating the Smartphone Addiction Scale Short Version (SAS-SV). Bivariate and multivariate analyses were performed using modified Poisson regression.

**Results:** The prevalence of smartphone addiction was 76.7% (95% CI: 71.4 to 81.2). Female students were 1.16 times more likely to be addicted than male students (APR: 1.16; 95% CI: 1.04 to 1.32). Students who spent more time on smartphones than on academic revision were 1.33 times more likely to be addicted (95% CI: 1.11 to 1.61). Those using smartphones for five or more hours daily were 1.32 times more likely to be addicted (95% CI: 1.02 to 1.48).

**Conclusion:** Smartphone addiction is highly prevalent at IUIU. Female gender and prolonged daily screen time are significant independent predictors. Targeted digital wellness programmes and institutional policy interventions are urgently needed.

## 1. INTRODUCTION

Smartphones have transformed daily life across all age groups. They provide access to communication, entertainment, education and social networking from a single handheld device. In the last decade the global smartphone user population has grown dramatically. By January 2021, approximately 5.22 billion people worldwide owned a smartphone, representing 66.6% of the global population (Datareportal, 2021). Within the same period, the number of users increased by 93 million in a single year.

University students represent one of the most active demographic groups in smartphone use. Academic applications, e-learning platforms and social media channels have made smartphones an integral part of student life (Ali et al., 2018). While these technologies offer clear educational benefits, excessive and uncontrolled use has given rise to behavioural patterns consistent with addiction, including loss of control, preoccupation and interference with daily functioning (Raza et al., 2020).

Smartphone addiction has been widely studied in Asia and Europe. Studies from Bangladesh found a prevalence of 61.4%, with male students showing higher rates (Zubair et al., 2022). Research in Saudi Arabia and Switzerland similarly reported high addiction rates among undergraduates (Alosaimi et al., 2016; Haug et al., 2015). In East Africa and sub-Saharan Africa broadly, systematic research on this topic remains sparse despite rapid smartphone penetration.

In Uganda, the Ministry of Education legalised smartphone use in secondary schools in 2021, making adolescent and young adult exposure to smartphones more pronounced. Yet no published study has rigorously examined smartphone addiction among Ugandan university students. This study sought to fill that gap by determining the prevalence of smartphone addiction and its associated sociodemographic and economic factors among students at the Islamic University in Uganda (IUIU) Kampala campus.

## 2. METHODS

### 2.1 Study Design and Setting

A cross-sectional study was conducted between April and August 2023 at IUIU Kampala campus. The campus is located on Kibuli Hill, approximately 3.5 kilometres south-east of Kampala central business district, and accommodates approximately 4,000 students across multiple faculties.

### 2.2 Study Population and Sampling

The target population comprised all undergraduate students enrolled at IUIU Kampala campus at the time of the study. Using Yamane’s formula with a 95% confidence level and a 5% margin of error, a sample size of 364 students was calculated. After accounting for non-response, 287 students were ultimately included in the final analysis. A simple random sampling technique was used to select participants who met the eligibility criteria.

Inclusion criteria: All students aged 18 years and above, enrolled in any programme at IUIU Kampala campus and in possession of a smartphone who gave written informed consent were included.

Exclusion criteria: Students on academic leave and those without a smartphone at the time of data collection were excluded.

### 2.3 Data Collection Instrument

Data were collected using a structured self-administered questionnaire divided into four sections. Section A covered sociodemographic data including age, gender, marital status, level of study and nationality. Section B examined academic performance indicators such as daily smartphone use duration, class attendance and examination retakes. Section C addressed economic factors including employment status and source of income. Section D used the validated Smartphone Addiction Scale Short Version (SAS-SV), a 10-item instrument scored on a 6-point Likert scale (Kwon et al., 2013). Students scoring above the established cut-off threshold were classified as smartphone-addicted.

### 2.4 Data Management and Analysis

Completed questionnaires were reviewed for completeness and coded. Data were entered into SPSS version 26 for analysis. Descriptive statistics were generated for all study variables. Chi-square tests were used in bivariate analysis to assess associations between predictor variables and smartphone addiction. Variables with a p value of less than 0.05 in bivariate analysis were entered into a modified Poisson regression model to obtain prevalence ratios (PR) with 95% confidence intervals.

### 2.5 Ethical Considerations

Ethical approval was obtained from the Faculty of Health Sciences of Islamic University in Uganda. Written informed consent was obtained from all participants prior to enrolment. Participation was voluntary. Data were anonymised using participant identification numbers to ensure confidentiality throughout the study.

## 3. RESULTS

### 3.1 Sociodemographic Characteristics

A total of 287 students participated in the study. The majority, 207 (72.4%), were aged between 18 and 24 years. More than half of the respondents, 154 (53.7%), were female. Most participants, 260 (90.6%), were single. Local students accounted for 197 (68.9%) of the sample, while the remaining 89 (31.1%) were international students. Regarding employment, 244 (85.0%) were not employed. Table 1 presents the full distribution of sociodemographic characteristics.

**Table 1:** Sociodemographic and economic characteristics of study participants (n=287)

| Variable | Frequency (n) | Percentage (%) |
| --- | --- | --- |
| **Age group (years)** |  |  |
| 18 to 24 | 207 | 72.4 |
| 25 to 31 | 42 | 14.5 |
| 32 to 38 | 30 | 10.5 |
| 39 and above | 8 | 2.6 |
| **Gender** |  |  |
| Male | 133 | 46.3 |
| Female | 154 | 53.7 |
| **Marital status** |  |  |
| Single | 260 | 90.6 |
| Married | 27 | 9.4 |
| **Nationality** |  |  |
| Local students | 197 | 68.9 |
| International students | 89 | 31.1 |
| **Level of study** |  |  |
| Year one | 49 | 17.1 |
| Year two | 47 | 16.4 |
| Year three | 79 | 27.5 |
| Year four | 55 | 19.2 |
| Year five | 57 | 19.9 |
| **Employment status** |  |  |
| Employed | 43 | 15.0 |
| Not employed | 244 | 85.0 |

### 3.2 Prevalence of Smartphone Addiction

The overall prevalence of smartphone addiction was **76.7%** (95% CI: 71.4 to 81.2), with 220 out of 287 students classified as addicted based on SAS-SV scores.

### 3.3 Bivariate Analysis

Table 2 shows the bivariate associations between sociodemographic factors and smartphone addiction. Gender and marital status were statistically significantly associated with addiction. Female students showed a higher addiction rate (81.8%) compared to male students (70.7%), with a p value of 0.026. Single students had a higher addiction rate (78.5%) compared to married students (59.2%), with a p value of 0.025. Age, nationality and employment status were not statistically significant.

**Table 2:**
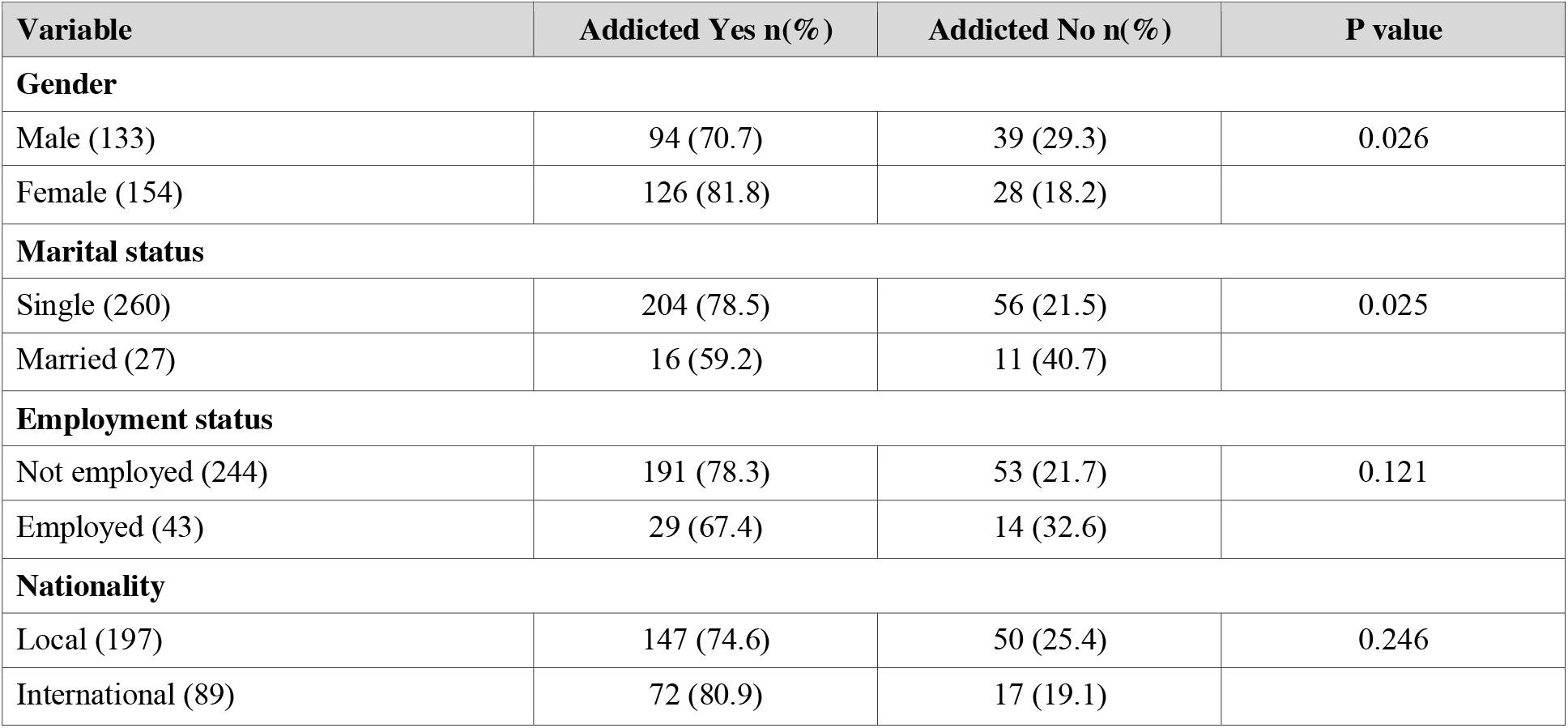
Bivariate analysis of sociodemographic and economic factors with smartphone addiction.

| Variable | Addicted Yes n(%) | Addicted No n(%) | P value |
| --- | --- | --- | --- |
| Gender |  |  |  |
| Male (133) | 94 (70.7) | 39 (29.3) | 0.026 |
| Female (154) | 126 (81.8) | 28 (18.2) |  |
| Marital status |  |  |  |
| Single (260) | 204 (78.5) | 56 (21.5) | 0.025 |
| Married (27) | 16 (59.2) | 11 (40.7) |  |
| Employment status |  |  |  |
| Not employed (244) | 191 (78.3) | 53 (21.7) | 0.121 |
| Employed (43) | 29 (67.4) | 14 (32.6) |  |
| Nationality |  |  |  |
| Local (197) | 147 (74.6) | 50 (25.4) | 0.246 |
| International (89) | 72 (80.9) | 17 (19.1) |  |

Table 3 presents the association between smartphone usage behaviours and addiction. Students who spent five or more hours daily on their smartphone had a significantly higher addiction rate of 81.9% compared to 64.9% among those who used it for one to four hours (p < 0.001). Students who reported spending more time on their smartphone than on academic revision had an addiction rate of 82.2% versus 63.1% (p < 0.001). Students who reported that smartphone use had affected their academic performance were also significantly more likely to be addicted (p = 0.039).

**Table 3:** Bivariate analysis of smartphone usage behaviours with addiction.

| Variable | Addicted Yes n(%) | Addicted No n(%) | P value |
| --- | --- | --- | --- |
| Daily screen time |  |  |  |
| 1 to 4 hours per day (77) | 50 (64.9) | 27 (35.1) | <0.001 |
| 5 hours and above (210) | 172 (81.9) | 38 (18.1) |  |
| More time on phone than revision |  |  |  |
| Yes (203) | 167 (82.2) | 36 (17.7) | <0.001 |
| No (84) | 53 (63.1) | 31 (36.9) |  |
| Smartphone affected class performance |  |  |  |
| Yes (105) | 92 (87.6) | 13 (12.4) | 0.039 |
| No (98) | 75 (76.5) | 23 (23.4) |  |

### 3.4 Multivariate Analysis

After adjusting for confounders in the modified Poisson regression model, three variables remained independently associated with smartphone addiction. Female students were 1.16 times more likely to be addicted compared to male students (APR: 1.16; 95% CI: 1.04 to 1.32; p = 0.031). Students who spent more time on their smartphone than on academic revision were 1.33 times more likely to be addicted (APR: 1.33; 95% CI: 1.11 to 1.61; p = 0.003). Students who used their smartphone for five or more hours per day were 1.32 times more likely to be addicted than those using it for fewer hours (APR: 1.32; 95% CI: 1.02 to 1.48; p = 0.034). These findings are summarised in Table 4.

**Table 4:**
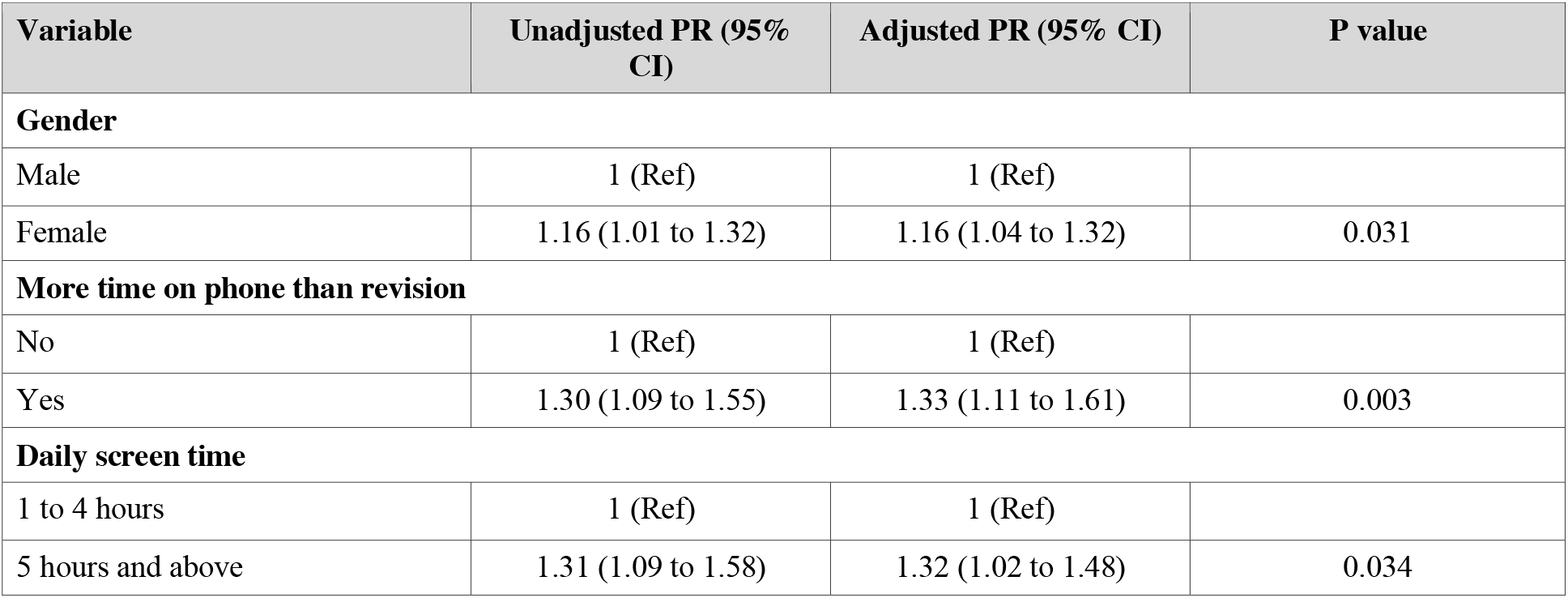
Multivariate analysis of factors associated with smartphone addiction (Modified Poisson Regression)

## 4. DISCUSSION

This study found a smartphone addiction prevalence of 76.7% among undergraduate students at IUIU, which is notably high compared to findings from other regions. A study conducted in Bangladesh reported a prevalence of 61.4% (Zubair et al., 2022), while studies in Saudi Arabia found rates ranging from 48% to 67% (Alosaimi et al., 2016). The higher rate observed in this study may reflect the growing dependency on smartphones for both social interaction and academic activity in a context where alternative digital infrastructure remains limited.

Female students were significantly more likely to be addicted in this study, consistent with findings from Saudi Arabia and across several East Asian settings (Lee, 2017). Researchers have attributed this pattern to higher engagement with social network services among young women, including greater use of messaging applications and social media platforms for relationship maintenance (Hakoama and Hakoyama, 2011). This finding underscores the need for gender-sensitive digital wellness interventions.

The strong association between prolonged daily screen time and addiction is consistent with existing literature. Students who used smartphones for five or more hours daily were significantly more likely to be classified as addicted. This threshold aligns with previous research that identifies daily duration of use as one of the most reliable behavioural predictors of addiction severity (Kahyaoglu-Sut et al., 2016). The finding that spending more time on smartphones than on academic revision independently predicted addiction highlights the direct academic cost of excessive smartphone use.

The high proportion of single students who were addicted (78.5%) may be explained by the greater role smartphones play in meeting social and emotional needs for unmarried young adults who have not yet established long-term social support structures through family life. This is consistent with findings from Bangladesh (Zubair et al., 2022).

Economic factors did not achieve statistical significance in multivariate analysis in this study, though studies from other settings have reported that students from higher income backgrounds show greater addiction rates due to access to more advanced devices and data (Zencirci et al., 2018). This difference may reflect the more homogenous socioeconomic background of students at IUIU relative to national universities in wealthier settings.

### Limitations

This study has several limitations. The cross-sectional design limits causal inference. The sample was restricted to one campus of IUIU and may not represent all Ugandan university students. Self-reported data may be subject to social desirability bias. Additionally, the SAS-SV was not originally validated in a Ugandan population, which may affect its measurement properties in this context.

## 5. CONCLUSION

Smartphone addiction is highly prevalent among undergraduate students at IUIU, with three in four students meeting the diagnostic threshold. Female gender and excessive daily screen time are the most significant independent predictors. Universities in Uganda and the wider East African region should develop evidence-based digital wellness programmes targeting at-risk student populations. Institutional policies regulating smartphone use in academic spaces, alongside student counselling initiatives, are warranted.

## ACKNOWLEDGEMENTS

The authors acknowledge the students of Islamic University in Uganda who voluntarily participated in this study, and the staff of Habib Medical School for their institutional support. Supervision by Dr. Aremu Babatunde Mujeeb is gratefully recognised.

## Competing Interests

All authors declare that they have no competing interests. No author or affiliated institution received any payment or services from a third party that could be perceived to influence this work.

## Funding

This study received no external funding. All costs were covered by the research team. No third party provided financial or material support for any aspect of the work.

## Data Availability

The dataset supporting the findings of this study is not publicly deposited but is available from the corresponding author, Mayanja Akram Mukalazi, upon reasonable request and subject to institutional data sharing guidelines of Islamic University in Uganda.

## Authors Contributions

MAM and AAA conceived and designed the study. BJ, AQK, HA, AS and AAS contributed to data collection and entry. MAM led the analysis and drafted the manuscript. All authors reviewed and approved the final version.

## Notes

### Competing Interest Statement

The authors have declared no competing interest.

### Funding Statement

This study did not receive any funding

### Author Declarations

Research and Ethics Committee (REC), Faculty of Health Sciences, Islamic University in Uganda (IUIU), Kampala Campus, Kibuli, Kampala, Uganda. Ethical approval was obtained and granted by the above committee prior to data collection.

